# The bladder microbiome of chronic kidney disease with associations to demographics, renal function, and serum cytokines

**DOI:** 10.1101/2023.05.05.23289568

**Authors:** Fengping Liu, Jingjie Du, Hao Lin, Zhenyi Xu, Yifan Sun, Feng Yan, Yifeng Gu, Yang Wang, Wei Guo, Qixiao Zhai, Jialin Hu, Yu Tian, Lei Hu, Peng Jiang, Shichao Wei, Chaoqun Gu, Jiayi Sheng, Wei Chen, Alan J. Wolfe, Ninghan Feng

**Author notes:** authors contributed equally to this article (Fengping Liu, Jingjie Du, Hao Lin, Zhenyi Xu, Yifan Sun, Feng Yan). **Correspondences: Wei Chen**. State Key Laboratory of Food Science and Technology and School of Food Science and Technology, Jiangnan University, Wuxi, Jiangsu, China 214122, **Alan J. Wolfe**. Department of Microbiology and Immunology, Stritch School of Medicine, Loyola University Chicago, Maywood, Illinois, USA 60153, **Ninghan Feng**. Department of Urology, Affiliated Wuxi No. 2 Hospital, Nanjing Medical University, Wuxi, Jiangsu, China 21400.

## Abstract

**Background:** High throughput 16S rRNA gene sequencing and enhanced culture methods (e.g., expanded quantitative urine culture, EQUC) have established the existence of the human bladder microbiome. We aim to test the hypothesis that the bladder environment of patients with chronic kidney disease (CKD) differs from that of unaffected controls with associated consequences for the composition of the bladder microbiome.

**Methods and Materials:** Females (n=66) and males (n=66) diagnosed with CKD and age-BMI-matched healthy control (HC) females (n=22) and males (n=22) were recruited. Transurethral catheterized urine was collected for 16S rRNA gene sequencing and Expanded Quantitative Urine Culture (EQUC). Fecal samples also were sequenced. Urinary analysis, kidney function and serum cytokines were examined.

**Results:** Bladder microbiomes of CKD females and males versus HC females and males differed (FDR<0.05); however, the difference was more obvious in females. In CKD females, sequencing revealed depletion of 5 genera, including *Lactobacillus*, and enrichment of 14 genera, including *Escherichia/Shigella, Bifidobacterium*, and several clostridial genera (FDR<0.05), while EQUC detected increased *Escherichia* and decreased *Lactobacillus* CKDB(P<0.05). *Escherichia-Shigella* was positively, whereas *Lactobacillus* was negatively, associated with CKDB-female serum creatinine (r=0.285, P=0.020; r=-0.337, P=0.006, respectively). *Lactobacillus* was positively associated with eGFR (r=0.251, P=0.042). Some CKD-related serum cytokines were negatively associated with clostridial genera. In contrast, the fecal microbiomes of CKD and HC females and males did not significantly differ in bacterial diversity or composition. However, bladder and fecal microbiomes of CKD females resembled each other more than those of controls, as assessed by the Bray-Curtis Dissimilarity Index (FDR<0.05).

**Conclusions:** CKD bladder microbiomes were dysbiotic, which was more obvious in females. This dysbiosis was associated with kidney damage severity and dysregulation of serum cytokines. The increased similarity between bladder and fecal microbiomes of CKD females suggests possible “gut-leakage.”

## 1 Introduction

Chronic kidney disease (CKD) is a non-communicable disease that affects more than 10% of the general population worldwide[1]. It is a progressive disease that over time leads to irreversible nephron loss and kidney failure. However, CKD is uncured by current therapies, possibly due to its unclear pathology. Previous studies found that microbial dysbiosis in human gut and blood is associated with CKD[2, 3], and crosstalk between host and microbiome is pathophysiologically relevant to CKD[2, 3].

The human bladder harbors its own microbial community[4–8]. Most studies have explored associations between the microbiomes of urine samples obtained by transurethral catheter (bladder) or midstream void (urogenital) and urinary tract disorders, such as urinary stone disease[9], urinary tract infection (UTI)[10], and bladder cancer[11]. One study assessed the urogenital microbiome in CKD patients and found it to be extremely diverse with greater diversity associated with estimated glomerular filtration rate (eGFR). However, this study did not include controls and analyzed voided urine samples[12], which often contains microbes from both the urinary and genital tracts[13, 14].

Kidneys are a major site for elimination of many cytokines. The delicate equilibrium of proinflammatory cytokines and their inhibitors is dysregulated in CKD[15], and associated with gut dysbiosis in CKD[16, 17]. For example, the dysregulation of tumor necrosis factor (TNF-α) and interleukin-6 (IL-6) is associated with gut dysbiosis in CKD[16, 17].

With mounting evidence of an interaction between gut microbiome and kidney function, some researchers have proposed a gut-kidney axis[18, 19]. However, these researchers ignored the possible existence of microbiomes in the urinary tract[20, 21]. Given that microbes are exquisitely sensitive to their environments and are known to interact with their niches, we hypothesize that (1) CKD patients have a dysbiotic bladder microbiome, (2) the bladder microbiome distinguishes CKD patients better than the fecal microbiome, and (3) the bladder microbiome is associated with serum cytokines.

## 2 Materials and methods

### 2.1 Study participant recruitment

Participants were recruited from July 1^st^ 2018 to March 30^th^ 2020. **Figure S1** displays the study design, including participant recruitment, methods, and objectives. Diagnostic criteria of CKD included decreased eGFR [<60 mL/min/1.73m^2^ or evidence of kidney damage, such as albuminuria (albumin excretion rate ≥30 mg/24 h; urinary albumin creatinine ratio [UACR] ≥30 mg/g], urine sediment abnormalities, electrolyte and other abnormalities due to tubular disorders, abnormalities detected by histology, and/or structural abnormalities detected by imaging or history of kidney transplantation[22]. CKD patients undergoing hemodialysis were excluded. Asymptomatic controls (“healthy” controls, HC) were recruited if they had an eGFR≥90 ml/min/1.73m^2^ and were free of current renal damage, a history of liver disease, cancer, diabetes, hypertension, and hyperlipidemia. After recruitment, HC subjects were excluded if they had an indicator of urinary tract infection (UTI) by urinalysis, such as >5 WBCs/HPF, >3 RBCs/HPF, positive urine nitrites, positive leucocyte esterase, positive occult, positive protein, and abnormal values of diabetes, hypertension, hyperlipidemia. Any participant with acute intercurrent disease or infection, diarrhea, kidney transplantation, pregnancy, breastfeeding was excluded. Those who used antibiotics, probiotics or immunosuppressive drugs within 30 days before enrollment also were excluded.

### 2.2 Urine collection and storage

Bladder urine samples were collected by transurethral catheterization and stored as reported previously[23]. Before catheter insertion, 5% iodophor was applied to disinfect the genital and perineal areas. Fifty mL urine were obtained and separated into three portions (i.e., 30 mL, 5 mL and 15 mL) that were used for 16S rRNA gene sequencing, Expanded Quantitative Urine Culture (EQUC) and clinical urinary analysis[4], respectively. All samples were processed in a biosafety cabinet. Samples for 16S rRNA gene sequencing and EQUC were placed in sterile, DNA- and enzyme-free centrifuge tubes. Samples for 16S rRNA gene sequencing were stored at -80°C refrigerator within 15 mins after collection. Samples for EQUC were placed in an insulated sterile container, immediately cooled on ice, and transferred to the research personnel within 4 hours.

### 2.3 Expanded Quantitative Bladder Urine Culture

Methods for EQUC were modified according to previous studies[6, 24]. 0.1 mL of catheterized bladder urine was inoculated onto BAP, chocolate and colistin, as well as nalidixic acid agars (Kemajia inc., Shanghai, China), streaked for quantification, and incubated in 5% CO_2_ at 35°C for 48h. Next, 0.1 mL of urine was inoculated onto two CDC anaerobe 5% sheep blood agar plates and incubated in an anaerobic incubator with a Campy gas mixture (5% O_2_, 10% CO_2_, 85% N_2_) at 35°C for 5 days. 1.0 mL of urine was placed in thioglycolate medium and incubated aerobically at 35°C for 5 days. If growth was visually detected in the thioglycolate medium, the medium was mixed, and 0.01 mL of urine was plated on BAP and CDC anaerobe 5% sheep blood agars for isolation and incubated aerobically and anaerobically at 35°C for 48h. Each morphologically distinct colony type was isolated on a different plate of the same media to prepare a pure culture that was used for identification, as described below.

### 2.4 16S rRNA gene sequencing

Bacterial DNA isolation from urine was described previously[6, 24]. The quantity and quality of the extracted DNA was measured using a NanoDrop ND-1000 spectrophotometer (Thermo Fisher Scientific, Waltham, MA, USA) and agarose gel electrophoresis. Using the isolated DNA as the template, the bacterial 16S rRNA V3-V4 region was PCR-amplified for 32 cycles using universal primers 338F and 806R. The resultant PCR amplicons were purified with Agencourt AMPure XP Beads (Beckman Coulter, Indianapolis, IN, USA) and quantified using the PicoGreen dsDNA Assay Kit (Invitrogen, Carlsbad, CA, USA). Amplicon pools were prepared for sequencing, and the size and quantity of the amplicon library were assessed using an Agilent 2100 Bioanalyzer (Agilent, Santa Clara, MA, USA) and Library Quantification Kit for Illumina (Kapa Biosciences, Woburn, MA, USA), respectively. The libraries were sequenced using the NovaSeq 6000 platform. As urines are low-biomass samples, they were sequenced in duplicate for reproducibility. When the Bray-Curtis dissimilarity of the duplicated urinary microbiome samples was above 0.3[25], they were re-sequenced. If their Bray-Curtis dissimilarity remained above 0.3[25], they were removed from further analysis. Six negative controls (2 without urine, 2 without bacterial DNA, and 2 without PCR product) also were sequenced.

### 2.5 Bioinformatic analysis of 16S rRNA gene sequencing data

Raw reads of the 16S rRNA gene sequences were trimmed using Cutadapt (cutadapt.readthedocs.io) to remove barcodes and adaptors. The overlapping paired-end reads were merged into a longer tag using FLASH (v1.2.8). Reads were quality trimmed using fdtrim (v0.94) from the 3’ end to remove bases with low-quality scores. Reads shorter than 100bp, with more than 5% Ns, or with an average quality below 20 were discarded. Chimeras were removed using Vsearch (v2.3.4). QIIME2 was used to process the clean reads to generate an ASV table, and taxonomy of microbes was identified using the Silva database (v138). Environmental contaminants of urine samples were removed as previously described[20]. Bacterial ASVs whose counts did not exceed five times the maximum number of counts in the negative controls were considered as contaminates and removed, as described[20]. We also manually removed bacteria that have been reported to be environmental contaminants from soil and water.

### 2.6 Identification of bacterial isolates

#### 2.6.1 16S rRNA gene sequencing of bacterial isolates

Bacterial isolates purified as described above were harvested: if the colonies were large enough, a single colony was chosen; if not, then the plate was scraped. To the harvested bacteria, 200μL lysis buffer (Guhe, Hangzhou, China) was added and its DNA extracted using the method described above for 16S rRNA gene sequencing. Identification of the isolates was confirmed by partial sequencing of the 16S rRNA gene. Briefly, the 16S rRNA gene was amplified for 30 cycles by PCR with universal primers 27F and 1492R, which results in an amplicon of approximately 1500 bp. PCR was performed in a DNA thermocycler (Thermo Scientific, USA) with PCR Premix (Ensure biologicals, Shanghai, China) in 50 μL reaction mixtures. The resultant amplicons were sequenced with the ABI Prism^TM^ Bigdye^TM^ terminator cycle sequencing reaction kit (Applied Biosystems, USA) using the 27F and 1492R primer pair. The sequencing product was purified using the Millipore-Montage dye removal kit (Ensure Biologicals, Shanghai, China) and sequenced on an ABI 3730XL capillary DNA sequencer (Applied Biosystems, USA). For each run, negative controls containing water instead of template DNA were run in parallel. All sequencing procedures were performed by BGI (Shanghai, China). For taxonomic identification, a portion of the sequence was compared to sequences in GenBank public database (http://www.ncbi.nlm.nih.gov/BLAST/).

#### 2.6.2 MALDI-TOF MS identification of bacterial isolates

When a bacterial isolate could not be identified by 16S rRNA gene sequencing, we used Matrix-Assisted Laser Desorption Ionization-Time-Of-Flight Mass Spectrometry (MALDI-TOF MS), as described previously[4]. Briefly, a small portion of a single colony was overlaid with 1 μL of a saturated solution of alpha-cyano-4-hydroxycinnamic acid matrix in 50% acetonitrile and 2.5% trifluoroacetic acid (VITEK MSCHCA, bioMérieux), then air dried. *Escherichia coli* (ATCC 8739) was used for system calibration. Mass spectra were acquired using a VITEK MS Plus (bioMérieux, Marcy l’Etoile) and the bioMérieux VITEK MS (IVD Knowledgebase v.3.2) used to analyze a composite mass spectrum for accurate identification. A probability score between 60% and 99.9% was considered to be a high discrimination value and thus a reliable identification. A probability score <60% was considered to be a low discrimination identification. No identification resulted when either no match was found for the composite spectra, or the analysis did not obtain enough spectral peaks[26]. Isolates that yielded no identification results were redeposited on the target plate and reanalyzed.

### 2.7 Other sample collection and processing

Blood samples for assessment of renal function and serum cytokines were collected on the day of urinary sample collection. The Bio-Plex^TM^ 200 System (Bio-Rad) and Bio-Plex Pro^TM^ Human Cytokine Screening 48-plex Panel (Bio-Rad, California, USA) were used to detect serum cytokines. An aliquot of bladder urine (described above) was used for urinalysis. The 24-hour urine volume was collected one day prior to the day of bladder microbiome sample collection. Fecal samples were collected on the day of bladder urine sample collection. The participants were told to excrete their feces into a sterile container. A nurse then put 30 mg feces into a sterile tube using sterile technique and stored them at -80°C refrigerator within 15 mins of collection. DNA isolation and sequencing of fecal samples was the same as for the urine samples, except that they were sequenced only once, as we have found that Bray-Curtis values for feces samples are typically <0.3.

### 2.8 Disease profiles assessment

Participants’ demographics data were collected during a face-to-face interview. Blood pressure and fasting blood glucose were assessed on the day of urine and fecal sample collection.

### 2.9 Statistical analysis

Pearson’s Chi-square or Fisher’s exact tests were used with categorical variables; Student’s *t* test was used on normalized continuous variables and Wilcoxon rank-sum test was used on non-normal continuous variables. The *P*-value was adjusted for multiple comparisons using the Benjamini–Hochberg (BH) false discovery rate (FDR). The significance threshold was set at an FDR-corrected value <0.05. Pearson correlation analysis was used to assess associations between variables, and the significance threshold was set at a P value <0.05.

## 3 Results

### 3.1 Participants

Figure **S1** displays the process, including participant recruitment, methods, and study objectives. Briefly, we collected urine samples through sterilized catheter from 230 CKD patients and 176 controls. All participants with catheterized urine (i.e., bladder) samples with bacterial DNA below the level of detection and control participants with abnormal urinalysis values were removed from downstream analysis. To ensure reproducibility, bladder urine samples were sequenced in duplicate, and participants whose samples were dissimilar (i.e., had a Bray-Curtis Dissimilarity score above 0.3) were removed from downstream analysis. From the remaining participants, age/BMI-matched CKD and HC males and females, resulting in 66 bladder urine samples from CKD males (CKDB-male) and 22 bladder samples from HC males (HCB-male) and 66 bladder samples from CKD females (CKDB-female) and 22 urine samples from HC females (HCB-female).

Compared to the HCB groups, both the male and female CKDB groups had higher rates of hypertension and diabetes, as well as higher rates of occult blood and cast in their urine (Fisher’s exact test, P<0.05; **Table 1**). Although both male and female CKDB groups had higher levels of systolic blood pressure than those in controls, only female patients had a higher level of diastolic blood pressure than that in controls (*t*-test, P<0.05; **Table 1**). Compared to HCB-males and HCB-females, the CKDB-males and CKDB-females had lower levels of eGFR and higher levels of serum creatinine, whereas the CKDB-males had a higher level of blood urea nitrogen and CKDB-females had a higher level of uric acid (*t*-test, P<0.05; **Table 1**). No difference was found in the nutrient intake between the groups of CKD and HC (*t*-test, P<0.05, **Table S1**); thus, it was not considered to be a confounding factor in downstream analyses.

**Table 1.** Demographics of urine donors of CKD patients and controls. Pearson Chi-square or Fisher’s exact test was used with categorical variables; Student’s t test on normalized continuous variables was used.

### 3.2 Bladder microbiomes are altered in CKD males and females

As gender difference has been reported to impact the bladder microbiome in asymptomatic subjects and disease groups[27, 28], we wondered whether the bladder microbiome composition of females and males differed between our CKDB and HCB groups. To test this, we performed principal-coordinate analysis (PCoA) of Bray-Curtis Dissimilarity indices for all microbial taxa (ASVs) present within and among the microbiomes of each group. As expected, the bladder microbiome of HCB-males and HCB-females was significantly separated (R^2^=0.077, FDR=0.002; **Figure 1A**); the bladder microbiome of CKDB-males and CKDB-females was also separated but less so (R^2^=0.017, FDR=0.016; **Figure 1A**). Since the bladder microbiome of males and females differed, further analyses were performed separately.

**Figure 1.**
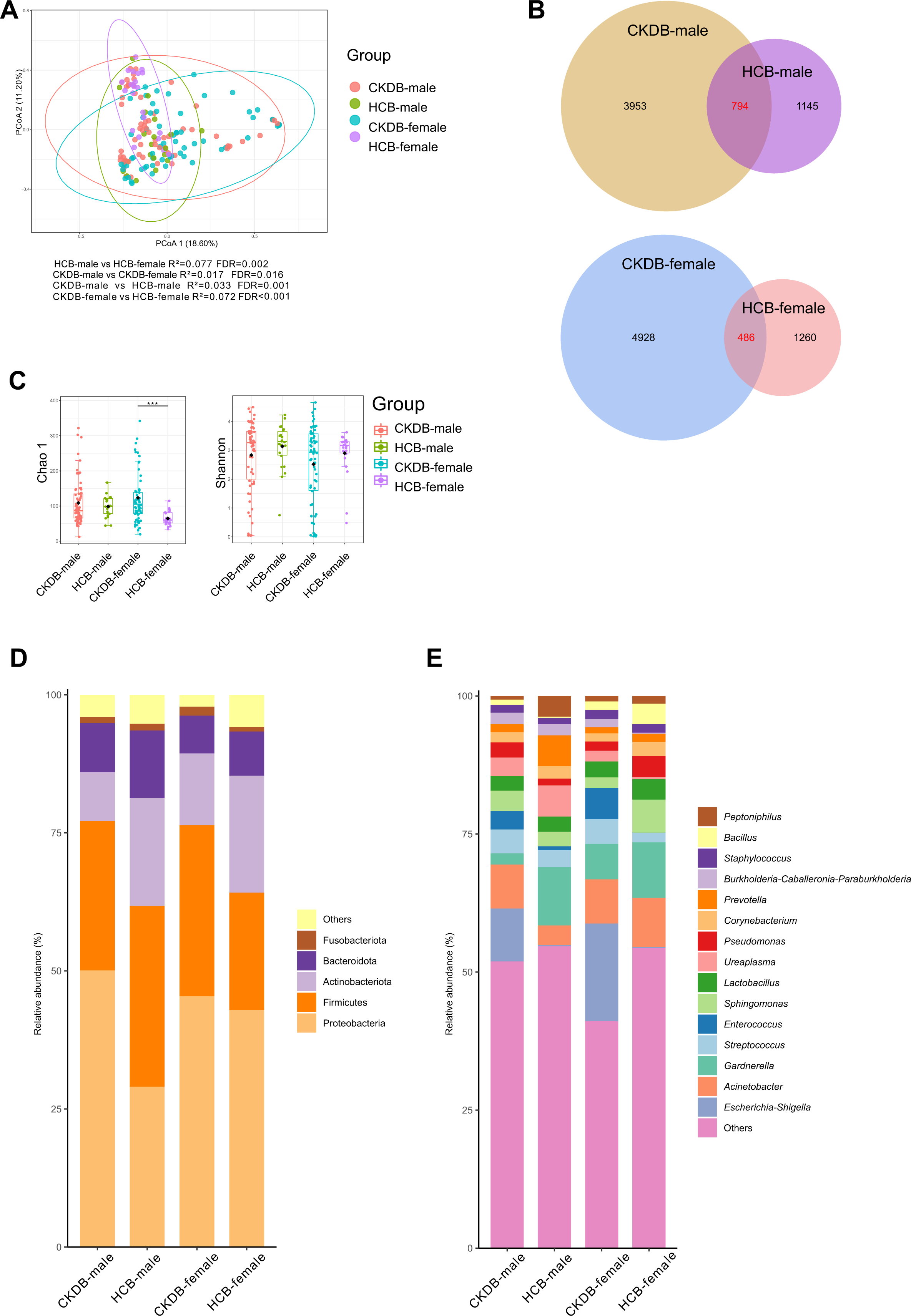
Bacterial composition, Venn, bacterial diversity, and phylum and genus composition in the urine from CKD patients and HC. **(A)** PCoA based on Bray-Curtis distances ASV level showed different microbial compositions between groups of CKDB-females and HCB-females, and between CKDB-males and HCB-males. The 95% confidence ellipse is drawn for each group. Permutational multivariate analysis of variance (PERMANOVA) was performed for statistical comparisons of samples in the two groups. *P* value was adjusted by the Benjamini and Hochberg false discovery rate (FDR). **(B)** Venn diagrams were made to compare the ASV composition within different groups. Each intersection of a number of areas represents the number of ASVs, which are present in all the intersecting areas. **(C)** Bacterial richness and diversity measured by Chao 1 and Shannon index were calculated at ASV level. Wilcoxon rank-sum test was performed and adjusted by Benjamini and Hochberg false discovery rate (FDR). ***, *P*_adj_ < 0.001. **(D)** Microbial profile at the phylum and genus level. Only the top 5 most abundant phylum and the top 15 most abundant genus are shown. Abrreviations: CKDB-female: urine samples provided by females with CKD; CKDB-male: urine samples provided by males with CKD; HCB-female: urine samples provided by healthy females; HCB-female: urine samples provided by healthy males.

When comparing CKDB to HCB, we found that the bladder microbiomes of both males and females differed (PERMANOVA, R^2^=0.033, FDR=0.001 and R^2^=0.072, FDR<0.001, respectively; **Figure 1A**). Hypertension, glucocorticoid levels, hypoglycemia and hypolipidemic agents were not confounding factors, as bladder microbiome of the medication users and gender-age matched non-users did not differ (PERMANOVA, FDR>0.05; **Figure S2A-D**). The bladder microbiome of CKDB-males and CKDB-females was significantly more diverse than those of their respective HCB controls. For females, this was reflected in both the number ASVs (**Figure 1B**) and the Chao 1 index, which estimate bacterial richness (Wilcoxon rank*-*sum test, FDR<0.001; **Figure 1C**); for males, this increased richness was reflected only in the number of ASVs (**Figure 1B**). To assess whether bacterial richness was associated with renal function in CKDB females, we performed a Pearson correlation analysis between Chao1 and estimators of renal function (i.e., eGFR, serum creatinine, blood urea nitrogen, and serum uric acid), but no association was observed (P>0.05).

### 3.3 Bladder microbiome composition is altered in CKD patients

Proteobacteria, Firmicutes, Actinobacteriota, Bacteroidota and Fusobacteriota were the 5 most abundant phyla in the bladder microbiomes of all 4 groups (**Figure 1D**). After correction for multiple testing, we observed no significant difference when we compared CKDB-males and HCB-males, and CKDB-females and HCB-females (Wilcoxon rank-sum test, FDR>0.05; **Table S2 & S3**). Notably, at the genus level, both the CKDB-males and CKDB-females were enriched for *Escherichia-Shigella* (9.59% and 17.71%, respectively), and *Enterococcus* (3.34% and 5.63% respectively; **Figure 1E**). Next, we compared bacterial genera that accounted for >0.1% of the total relative abundance in males and females. After correction for multiple testing, we observed no significant difference when we compared CKDB-males and HCB-males (Wilcoxon rank-sum test, FDR>0.05; **Table S4**). In contrast, 19 genera differed significantly between CKDB-females and HCB-females (Wilcoxon rank-sum test, FDR<0.05; **Table S5**). Five genera were significantly less abundant in CKDB-females relative to HCB-females (Wilcoxon rank-sum test, FDR<0.05). These included one unknown genus in the family Comamonadaceae and the genera *Lactobacillus, Sphingobium*, *Sphingomonas*, and *Streptomyces* (**Table S5**; **Figure 2A**). Fourteen genera were significantly more abundant in CKDB-females, including *Escherichia-Shigella*. Others included *Agathobacter*, *Anoxybacillus*, *Bacteroides*, *Bifidobacterium*, *Blautia*, *Comamonas*, *Diaphorobacter*, [Eubacterium] coprostanoligenes group, *Faecalibacterium*, CAG-352 in the family Ruminococcaceae, *Sphingobacterium*, and *Subdoligranulum* (Wilcoxon rank-sum test, FDR<0.05; **Table S5**; **Figure 2B**). Many of these taxa are members of the class Clostridia. As *Escherichia-Shigella* is one of most common causes of UTI[29, 30], and the participants with current UTI were excluded, we compared the abundance of *Escherichia-Shigella* between the age-matched CKDB-female urine samples with positive urinary leucocyte esterase and negative ones but there was no significant difference between the positive samples and negative ones (29.82±45.23 vs. 24.73±39.24 (Wilcoxon rank-sum test, P=0.781, FDR=0.814). In addition, since *Lactobacillus* can impede *Escherichia* growth in the urinary tract[31], we performed Pearson correlation analysis to examine whether they were connected but there was no significant correlation (r=-0.221, P=0.075).

**Figure 2.**
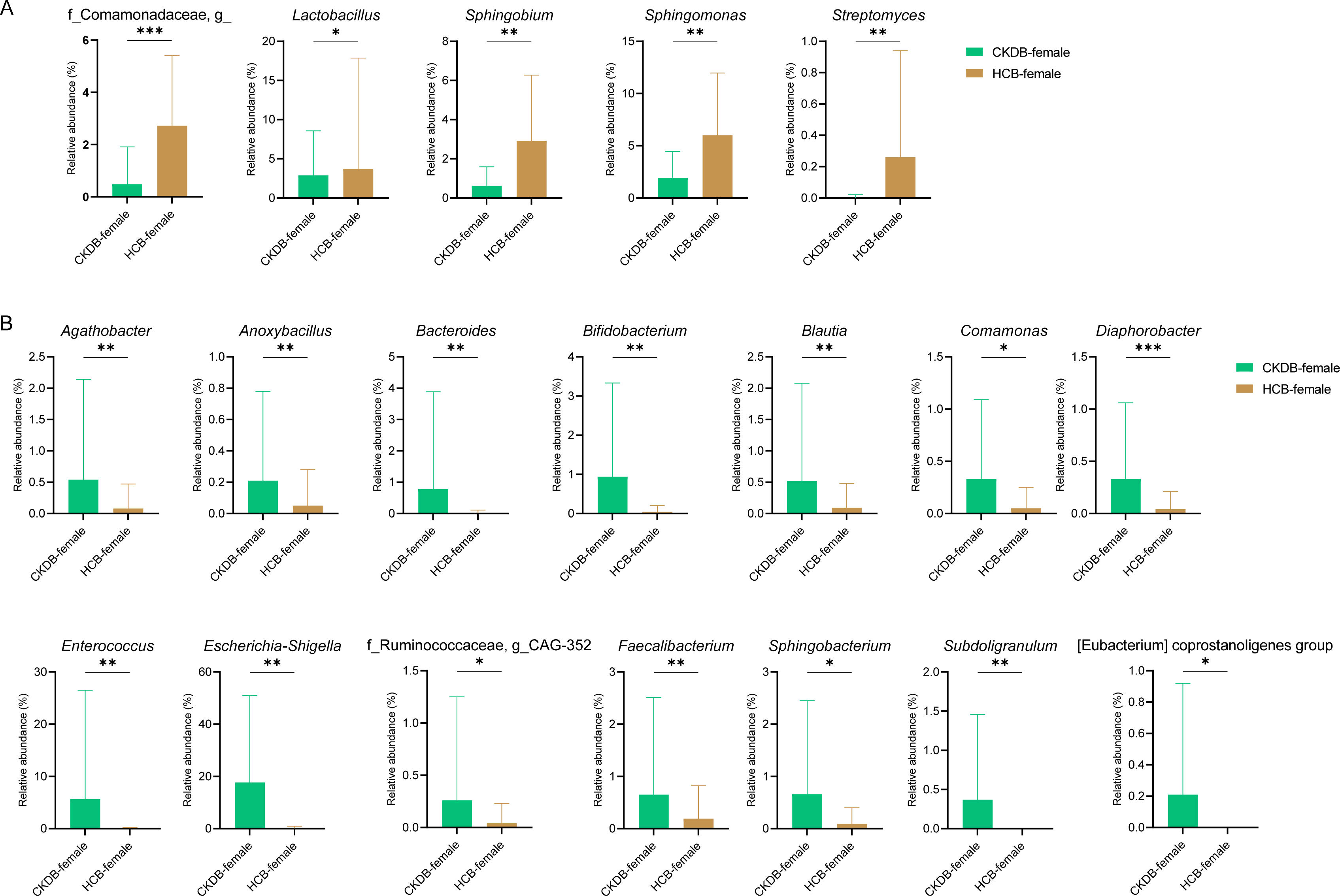
Bacterial genera that were differentially abundant between CKD patients and controls. **(A)** Bacterial genera significantly decreased in CKDB-females comparing to those in HCB-females. **(B)** Bacterial genera significantly increased in CKDB-females comparing to those in HCB-females. P value was calculated using Wilcoxon rank-sum test and adjusted by Benjamini and Hochberg FDR. *, FDR < 0.05; **, FDR<0.01; and ***, FDR< 0.001. Abrreviations: CKDB-female: urine samples provided by females with CKD; CKDB-male: urine samples provided by males with CKD; HCB-female: urine samples provided by healthy females; HCB-female: urine samples provided by healthy males.

To determine whether the detected sequences came from live bacteria, we performed EQUC (**Table S6**), an enhanced culture method that vastly outperforms the standard clinical microbiology urine culture method[4, 32]. Microbes (including both bacteria and fungi) were more prevalent in HCB-females than in HCB-males (68.2 vs 13.6%, P<0.001; **Table S7**), whereas microbes were only slightly more prevalent in CKDB-females than in CKDB-males (51.1 vs 42.4%, P=0.295; **Table S8**). Microbes also were more prevalent in CKDB-males than in HCB-males (42.2 vs. 13.6%, P=0.014; **Table S9**). In contrast, microbes were equally prevalent in CKDB-females and HCB-females (51.5 vs 68.2%, P=0.173; **Table S10**).

Only two identified genera and one unidentified genus were detected in HCB-males: the fungus *Cladosporium* was detected in one (4.5%), the bacterium *Curtobacterium* was detected in another (4.5%), and an unidentified genus was detected in a third (4.5%). In contrast, multiple genera were detected in CKDB-males. The most prevalent genera were *Bacillus* [18.18% (11/66)], *Staphylococcus* [9.09% (6/66)], *Kocuria* [3.03% (2/66)], *Micrococcus* [3.03% (2/66)], and *Paenibacillus* [3.03% (2/66)]. The genus *Bacillus* was significantly more prevalent in the CKDB-males relative to HCB-males (Fisher’s exact test, P=0.031; **Figure 3A**).

**Figure 3.**
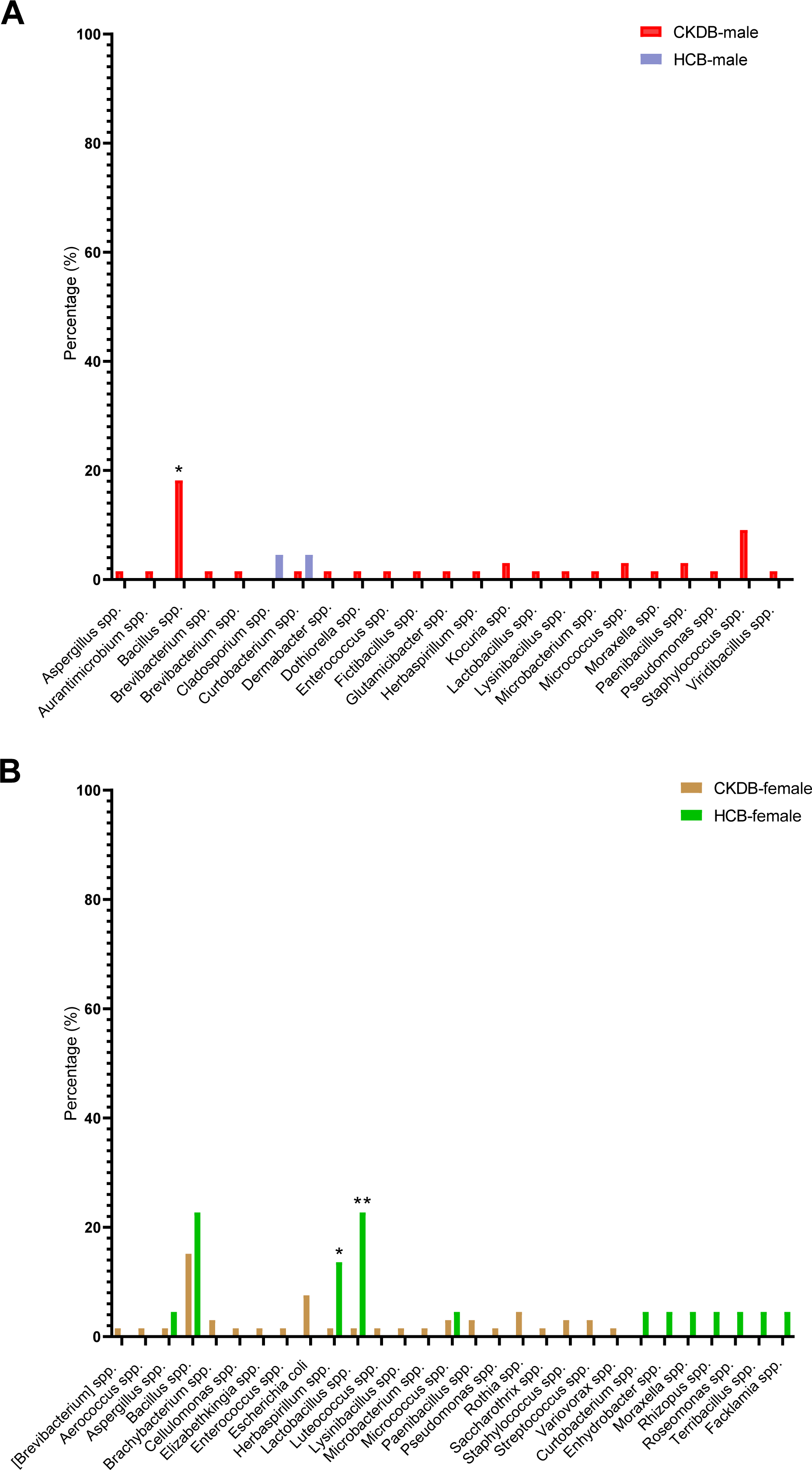
Comparisons of numbers of subjects with visible bacterial isolates between groups of patient and HC. Chi-square/Fisher’s test was used, P values that are significant at 0.05. Bars represent the prevalence of isolates in each group. Abrreviations: CKDB-female: urine samples provided by females with CKD; CKDB-male: urine samples provided by males with CKD; HCB-female: urine samples provided by healthy females; HCB-female: urine samples provided by healthy males.

Although the overall prevalence of microbes did not differ between HCB-females and CKDB-females, many genera differed (**Figure 3B**). The most prevalent in HCB-females were *Bacillus* [22.73% (5/22)], *Lactobacillus* [22.73% (5/22)], and *Herbaspirillum* [13.64% (3/22)], whereas the most prevalent in CKDB-females were *Bacillus* [15.15% (10/66)], *Escherichia* [7.58% (5/66)], and *Rothia spp*. [4.55% (3/66)]. The genera *Lactobacillus* and *Herbaspirillum* were significantly less prevalent in CKDB-females than in HCB-females (Fisher’s exact test, P<0.05; **Figure 3B**).

### 3.4 Altered genera in CKD was associated with demographics and renal function

Because the bladder microbiome composition of CKDB-females and HCB-females differed, we used Pearson correlation analysis to determine whether any of the significantly different taxa (as determined by 16S rRNA gene sequencing) were associated with any demographic category or renal function. For CKDB-females, multiple associations were observed (**Figure 4A**). For example, *Escherichia-Shigella* was positively associated with CKDB-female serum creatinine (r=0.285, P=0.020). *Lactobacillus* was negatively associated with CKDB-female age and serum creatinine (r=-0.330, P=0.007 and r=-0.337, P=0.006; respectively), but positively associated with CKDB-female eGFR (r=0.251, P=0.042). *Bifidobacterium* was positively associated with CKDB-female disease duration (r=0.335, P=0.006). In contrast, significant associations were not observed for HCB-females (r<0.03, P>0.05; **Figure 4B**), with one exception: *Streptomyces* was positively associated with blood urea nitrogen (r=0.790, P<0.001).

**Figure 4.**
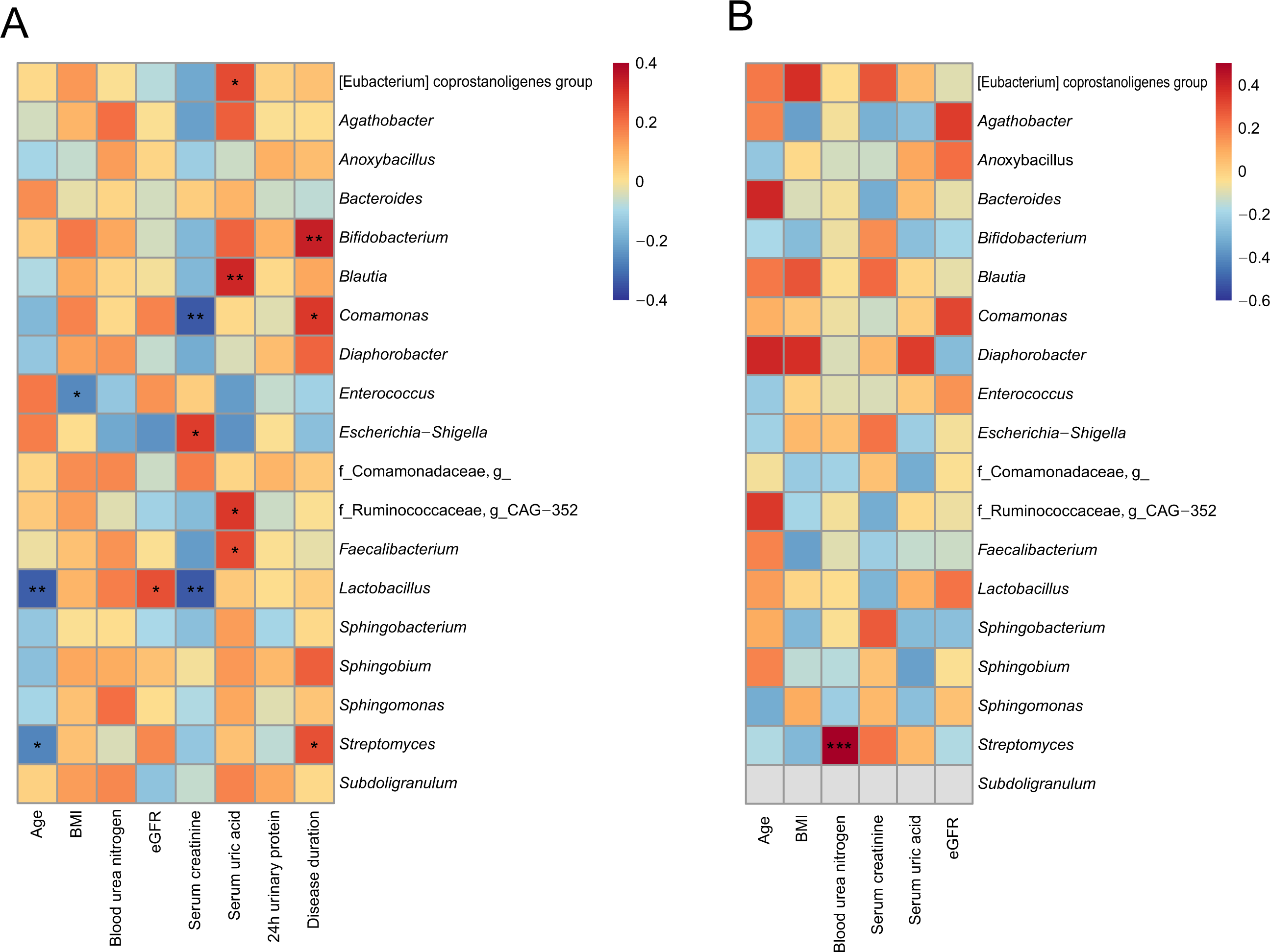
Bladder microbiome was associated with participants’ demographics and kidney function estimators. **(A)** The heatmap depicted the association between the bacterial genera showing differed in CKDB-female and CKD-female’s demographics and kidney function estimators. **(B)** The heatmap depicted the association between the bacterial genera showing differed in HCB-female and HC-female’s demographics and kidney function estimators. Pearson correlation analysis was performed. The correlation of two variables with values of |*r*|>0.3 and *P*□*<*□0.05 are displayed. *, *P*□*<*□0.05; **, *P*□*<*□0.01; and ***, *P*□*<*□0.001.

### 3.5 Bacterial community altered in all stages of CKD patients

For each CKD stage, we performed PCoA of Bray-Curtis Dissimilarity indices, comparing their bladder microbiomes to gender- and age-matched controls; these microbiomes differed significantly in stages 2-5 (PERMANOVA, FDR<0.05), but not stage 1 (PERMANOVA, FDR>0.05; **Figure S3A**). These differences were primarily due to changes in composition, as only CKDB microbiomes from stages 3 and 5 were significantly richer than their respective HCB (as measured by Chao 1; Wilcoxon rank*-*sum test, FDR<0.05) and the Shannon indices did not differ (**Figure S3B**).

### 3.6 Altered bladder microbiome associated with altered serum cytokines in CKD patients

To determine if associations exist between the bladder microbiome and serum cytokines, we first measured serum cytokines and then compared them to the bladder microbiome. Of the 48 serum cytokines we assessed, 44 cytokines were detected. Of these, 23 cytokines differed significantly between the 66 CKD females and 22 female controls (Wilcoxon rank-sum test, FDR<0.001). Some, such as IL-8, IL-18, IL-1β and TNF-α were increased in CKD females, whereas others, such as IL-9 and TNF-β were decreased in CKD females (**Figure 5A**). To determine whether associations existed between serum cytokines and bladder microbial composition, we performed Pearson correlation analysis. Several taxa enriched in HCB-females were negatively associated with increased serum cytokines (r>0.03, P<0.05; **Figure 5B**). For example, the genera *Sphingobium* and *Streptomyces* were negatively associated with IL-18, IL-8, and/or IL-1β. In contrast, several taxa enriched in CKDB-females were positively associated with increased levels of serum cytokines (r>0.03, P<0.05; **Figure 5B**). For example, *Bifidobacterium* was positively associated with IL−2Rα, and MIG (r>0.03, P<0.05), and IL-8 was positively associated with the members of Clostridia, including *Agathobacter*, *Blautia*, *Faecalibacterium*, *Subdoligranulum* (r>0.03, P<0.05). Intriguingly, the genus *Lactobacillus* was enriched in HCB-females but none of the correlations were statistically significant (**Figure 5B**).

**Figure 5.**
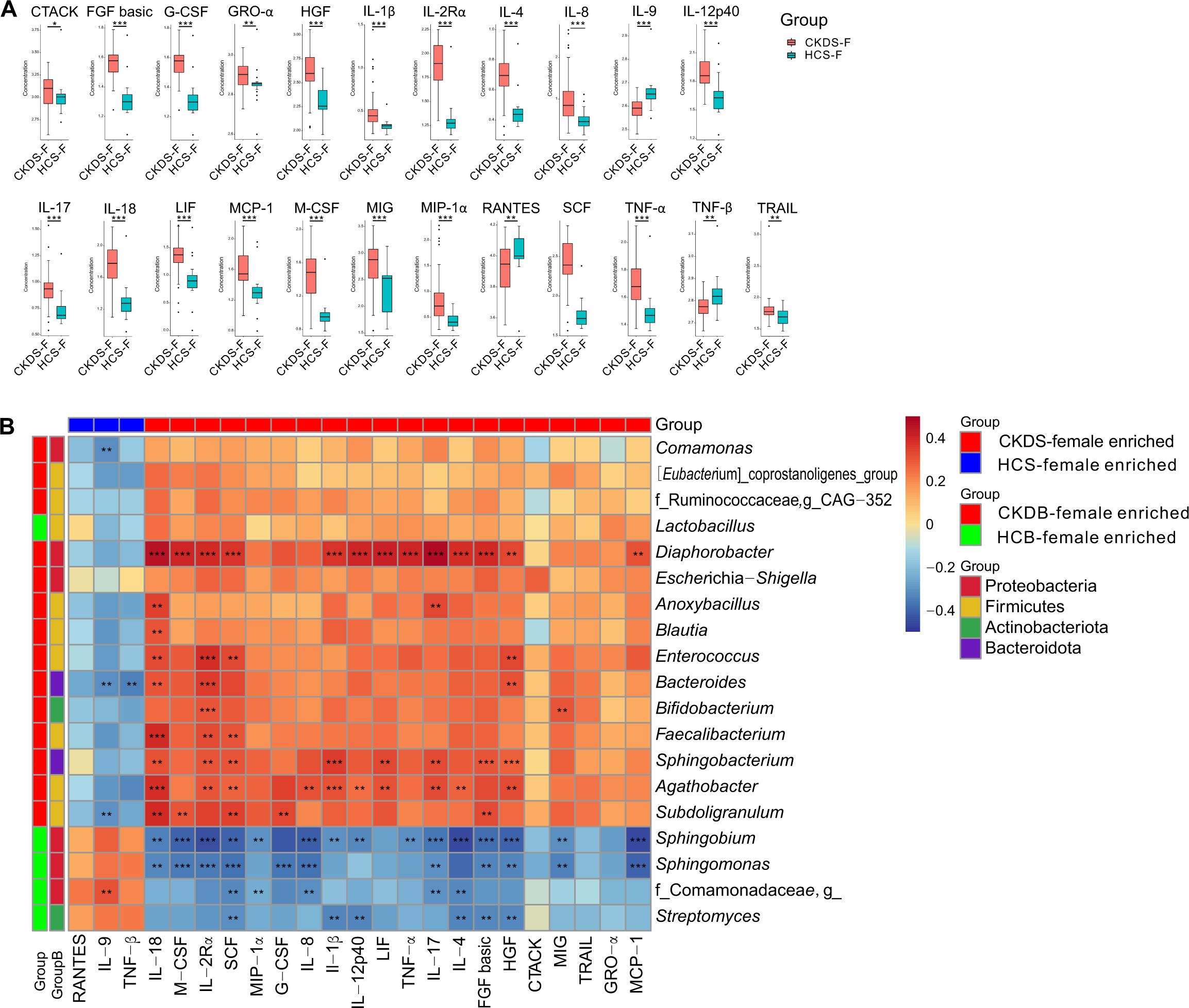
Serum cytokines profiles in CKD females and their associations to urinary microbiome. **(A)** Serum cytokines increased and decreased in CKD females compared to controls. *P* value was calculated using Wilcoxon rank-sum test and adjusted by Benjamini and Hochbergs false discovery rate (FDR). **(B)** Pearson correlation analysis was performed on the bacterial genera and cytokines that differed between the CKD females and controls. The correlation of two variables with values of |*r*|>0.3 and *P*□*<*□0.05 are displayed. *, *P*□*<*□0.05; **, *P*□*<*□0.01; and ***, *P*□*<*□0.001. Abrreviations: CKDB-female: urine samples provided by females with CKD; CKDS-female: serum samples provided by females with CKD; HCB-female: urine samples provided by healthy females; HCS-female: serum samples provided by healthy females.

### 3.7 Alteration of microbiome not observed in feces

To assess the gut-bladder axis in CKD, we assessed the fecal and bladder microbiomes of CKD-males (n=10) to age-matched HC-males (n=10) and CKD-females (n=22) with age-matched HC-females (n=22) who provided both fecal and catheterized urine samples. We observed no difference in gut microbiome compositions (PERMANOVA, FDR>0.05; **Figure 6A**) or alpha diversity (Wilcoxon rank-sum test, FDR>0.05; **Figure 6B**) between fecal samples from CKD males (CKDF-males) and fecal samples from HC males (HCF-males) or between fecal samples from CKD females (CKDF-females) and fecal samples from HC females (HCF-females). To test the hypothesis that gut-kidney leakage occurs in CKD patients, we first performed PCoA on the bladder and fecal microbiomes in CKD patients and controls; the bladder microbiomes differed significantly from the corresponding fecal microbiomes for both CKD-males and females and their age-matched HC-controls (PERMANOVA, FDR<0.05; **Figures 6C-6F**). We then compared the Bray-Curtis dissimilarity between the bladder and gut microbiomes from either CKD males/females or HC males/females. These microbiomes tended to be less dissimilar in CKD-females than in HC-females (Wilcoxon rank-sum test, FDR<0.001), but not in the males (Wilcoxon rank-sum test, FDR>0.05; **Figure 6G**). In addition, the Bray-Curtis dissimilarity between the bladder and gut microbiomes from CKD females was slightly less than that from CKD males (0.965±0.068 vs. 0.983±0.048, Wilcoxon rank-sum test, FDR=0.089), while the Bray-Curtis dissimilarity between the bladder and fecal microbiomes from healthy females was almost equal to that from healthy males (0.998±0.005 vs. 0.998±0.004, Wilcoxon rank-sum test, FDR=0.472).

**Figure 6.**
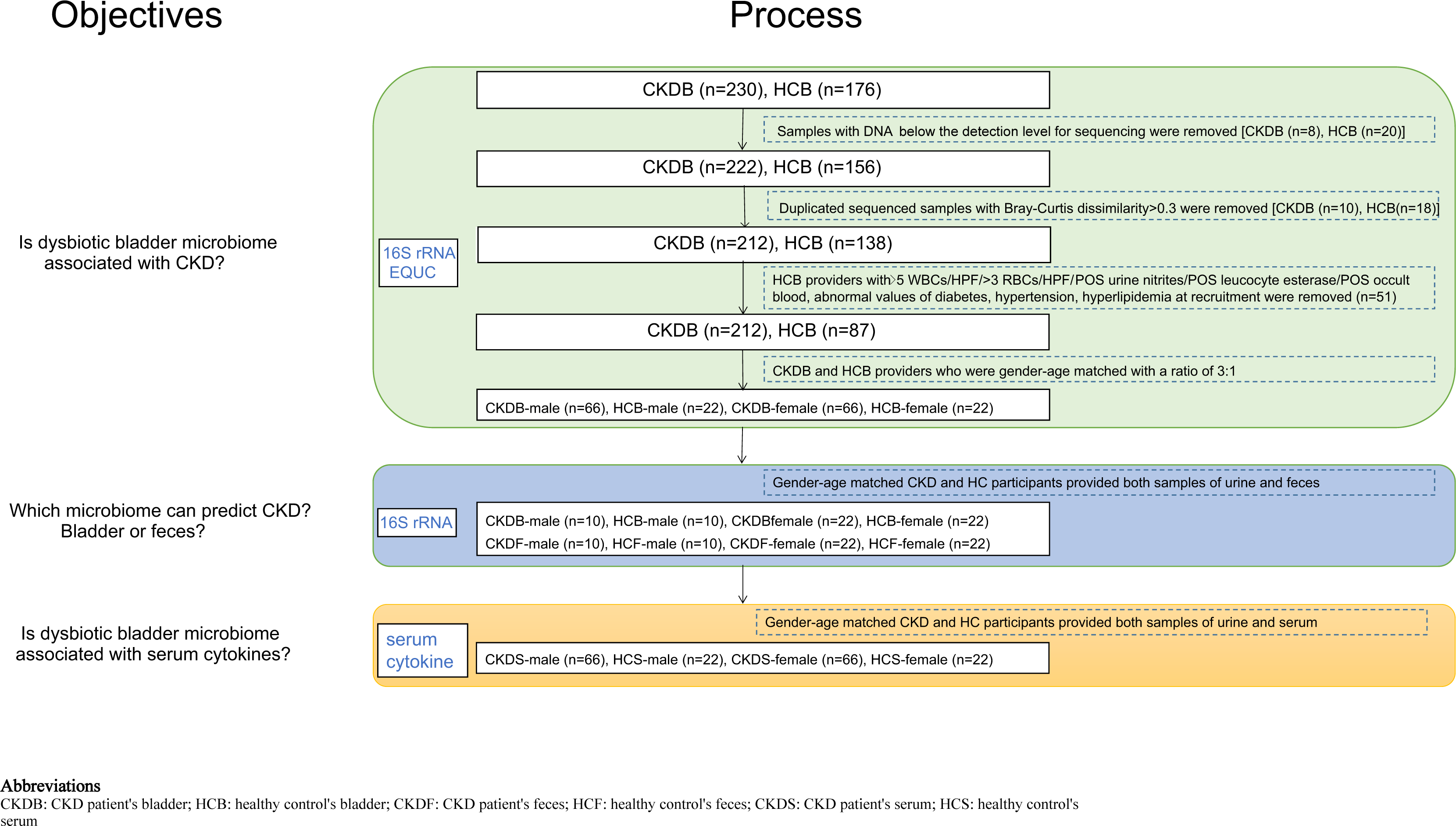
Comparisons of microbiomes in feces and urine in CKD males and females. **(A)** PCoA based on Bray-Curtis distances ASV level showed different microbial compositions between groups of CKDF-females and HCF-females, and between CKDF-males and HCF-males. The 95% confidence ellipse is drawn for each group. Permutational multivariate analysis of variance (PERMANOVA) was performed for statistical comparisons of samples in the two groups. *P* value was adjusted by the Benjamini and Hochberg false discovery rate (FDR). **(B)** Bacterial richness and diversity measured by Chao 1 and Shannon index were calculated at ASV level. Wilcoxon rank-sum test was performed and adjusted by Benjamini and Hochberg false discovery rate (FDR). **(C-F)** PCoA based on Bray-Curtis distances ASV level showed different microbial compositions between groups of CKDF-male and CKDB-male; between HCF-males and HCB-males; between CKDF-female and CKDB-female; between HCF-female and HCB-female. The 95% confidence ellipse is drawn for each group. Permutational multivariate analysis of variance (PERMANOVA) was performed for statistical comparisons of samples in the two groups. *P* value was adjusted by the Benjamini and Hochberg false discovery rate (FDR). **(G)** Comparisons of dissimilarity of Bray-Curtis between females and males’ urine and feces in patients and controls group. Wilcoxon rank-sum test was performed and adjusted by Benjamini and Hochberg false discovery rate (FDR). **, FDR < 0.01. Abrreviations: CKDF-female: fecal samples provided by females with CKD; CKDF-male: fecal samples provided by males with CKD; CKDB-female: urine samples provided by males with CKD; CKDB-male: urine samples provided by males with CKD; HCF-female: fecal samples provided by females with CKD; HCF-male: fecal samples provided by males with CKD; HCB-female: urine samples provided by healthy females; HCB-male: urine samples provided by healthy males.

#### Discussion

The role of a balanced intestinal microbiome in maintaining kidney function is widely recognized[2]; however, the role of the bladder microbiome in kidney functions remains understudied. This study is the first to reveal that CKD patients possess a distinct bladder microbiome, and that the bladder microbial profile is associated with unbalanced kidney function and dysregulated serum cytokine profiles.

First, we found that both male and female CKD patients had dysbiotic bladder microbiomes, as assessed by 16S rRNA gene sequencing. This dysbiosis may be associated with loss of kidney function, as the bladder microbiomes of CKD patients in stages 2-5 differed from those of controls. A similar phenomenon in the gut was described by Wu et al., who reported that the fecal microbiome could reflect the CKD stage[33].

We observed an increased microbial richness in CKD patients relative to controls and found that this increase was greater in females than in males, as assessed by both sequencing and EQUC. Since population-based studies show that CKD affects more females than males[34], bladder dysbiosis in the context of CKD sex differences is worthy of future exploration.

One striking finding of this study is that CKD females’ bladder microbiome was enriched for the genus *Escherichia*/*Shigella*, as assessed by both 16S rRNA gene sequencing and EQUC. This genus was not associated with urinary leucocyte esterase or serum cytokine profile, but its increase in CKD females was positively correlated to the level of serum creatinine, a kidney function estimator. Given that *Escherichia/Shigella* is the most commonly detected genus in UTI[29, 30], its increased prevalence at relatively high abundance in CKD females indicate that this potentially uropathogenic microbe should be a concern during treatment of CKD.

Another notable finding of this study is that both detection methods (sequencing and EQUC) revealed a depletion of *Lactobacillus* in CKD females. Decreased *Lactobacillus* has been reported in patients with UTI[35], and may be a sign of an unhealthy bladder. Even though we did not observe an association between *Lactobacillus* and serum cytokines like previous urinary microbiome studies on interstitial cystitis and systemic lupus erythematosus suggested[20, 36], *Lactobacillus* was associated with the kidney function indicators. It was positively associated with eGFR and negatively associated with serum creatinine, indicating that *Lactobacillus* in the bladder may be a biomarker for kidney function. Although the effects of bladder *Lactobacillus* on kidney functions have never been reported, *Lactobacillus* oral supplementation in mouse models has been suggested to attenuate the loss of kidney function, in a process that involved inflammation pathways[37, 38]. Thus, *Lactobacillus* in the bladder might protect kidney function in females.

While decreased *Lactobacillus* in the vagina is a well-documented feature of older females, recent studies have shown this decrease to also be true of the bladder. Komesu et al. found that the relative abundance of *Lactobacillus* in catheterized urine had a strong negative relationship with age when study participants with mixed urinary incontinence and controls were assessed together[39]. Another study demonstrated that *Lactobacillus* in mid-stream voided urine was more common in pre-menopausal females comparing to post-menopausal ones[40]. In contrast, our study found a negative association between bladder *Lactobacillus* and age only in female CKD patients. This finding suggests that loss of kidney function, a sign of getting older[41], may be attributable to *Lactobacillus* depletion.

*Bifidobacterium* is widely advocated as a probiotic due to its potential beneficial effects[42]. However, in our study, bladder *Bifidobacterium* was enriched in CKD females and positively associated with disease duration and MIG, a chemoattractant cytokine for activated T lymphocytes and tumor infiltrating T-lymphocytes[43], and an indication of an unhealthy kidney, e.g. kidney cancer or stones[44, 45]. Increased *Bifidobacterium* also has been reported in patients with kidney stones[21]. *Bifidobacterium* has been reported in bladder infections and other diseases [46–48]. Therefore, it is unclear whether *Bifidobacterium* in the bladder serves as a probiotic or not.

The upregulation of IL-1β, IL-8, IL-18 and TNF-α in CKD patients have been reported[49–51]. As expected, all these cytokines were upregulated in the serum of CKD females, and almost all were negatively associated with decreased *Sphingobium* in CKD female bladders. As *Sphingobium* has been rarely reported, the roles of these associations remain to be examined in the future. It is worth noting that half of the bacterial genera enriched in CKD females are members of the class Clostridia, including *Agathobacter*, *Blautia*, *Faecalibacterium* and *Subdoligranulum*, and all were positively linked to the increase of the level of IL-18 in patients. Some members of Clostridia, including Clostridiales, Lachnospiraceae and Ruminococcaceae, have been reported to be enriched in the voided urine of patients with chronic prostatitis[52]. It is reasonable to conclude that Clostridia plays a pathogenic role in CKD females.

To explore the gut-kidney axis, we assessed the fecal and bladder microbiomes in age-gender-matched CKD patients and controls. We only observed a statistical difference in the bladder microbiome. Using the Bray-Curtis Dissimilarity Index, we compared the fecal and bladder microbiomes and observed a statistically significant decrease in the female CKD patients but not in the matched controls, supporting the hypothesis that “gut-leakage” may occur in CKD females.

Our study has limitations. First, the number of CKD patients and age-gender-BMI matched controls were unequal with a 3:1 patient:control ratio. Thus, there may exist additional confounders that vary within the patients and controls[53]. However, kidney-age-related problems are one of the most common health issues in elderly population[41], so it was difficult to recruit healthy matched controls. Second, we only enrolled from a single institution which cannot appropriately power the genetic profiles[54].

In summary, our study shows that the bladder microbiome of patients with CKD differs significantly from that of the controls, especially in females. The bladder microbiomes of CKD patients are richer (i.e., include more taxa) and many of those taxa are distinctly different in prevalence and abundance between two groups. Both 16S rRNA gene sequencing and EQUC detected a significant increase of *Escherichia*/*Shigella* and a significant decline of *Lactobacillus* in CKD females, and such alterations may be related to reduced kidney function. We observed associations between several bacterial genera in the bladders of CKD patients with several CKD-associated serum cytokines, suggesting that these genera might play a pathological role. Lastly, bladder dysbiosis was accompanied by “gut-leakage” in CKD females. Further studies are needed to investigate how the bladder microbiome modifies kidney function, inflammation and “gut leakage” using CKD animal models.

## Data Availability

All data produced are available online at https://www.ncbi.nlm.nih.gov/bioproject/PRJNA807874

## Declarations

### Ethics approval and Consent to participate

The ethics committee of the Affiliated Wuxi Second Hospital of Nanjing Medical University approved the study (Ref. 2018051). Informed consent was provided by all subjects prior to sample collection.

### Consent for publication

All authors consent to the publication of this manuscript.

### Availability of data and materials

Raw data from 16S rRNA sequencing are available in the Sequence Read Archive under BioProject ID SRP360378 (https://www.ncbi.nlm.nih.gov/bioproject/PRJNA807874).

### Competing interests

All authors declare that they have no competing interests.

### Funding

Wuxi “Taihu Talents Program” Medical and Health High-level Talents Project (THRCJH20200901); Wuxi “key medical discipline construction” Municipal Clinical Medical Center (municipal public health center) Project (LCYXZX202103); Wuxi Technological Project (N20192047); Zhejiang Provincial Natural Science Foundation of China (LXR22H160001); National Natural Science Foundation of China (81874142 and 82073041).

### Authors’ contributions

Conceptualization: Fengping Liu, Alan J. Wolfe, Wei Chen, and Ninghan Feng; methodology: Fengping Liu, Jingjie Du, Qixiao Zhai, Hao Lin, Yifan Xu, Feng Yan, Yang Wang, Wei Guo, Zhenyi Xu, Jialin Hu, Peng Jiang, Chaoqun Gu, Jiayi Sheng, Lei Hu; software: Fengping Liu, and Jingjie Du; validation: Alan J. Wolfe; writing: Alan J. Wolfe, Fengping Liu, Jingjie Du, Yifeng Gu; supervision: Alan J. Wolfe; funding acquisition: Ninghan Feng; project administration: Fengping Liu, Jialin Hu, Lei Hu, Peng Jiang, Jiayi Sheng, Chaoqu Gu, Yang Wan, FengYan, Hao Lin, Shichao Wei.

## Acknowledgements

We gratefully acknowledge the volunteers who participated in our study.

## Legends

**Table S1** Comparison of nutrient intake between CKD and HC cohort.

Student’s *t* test on normalized continuous variables, and Wilcoxon rank-sum test on non-normal continuous variables.

**Tables S2** Comparison of bacterial phylum abundance between CKDB-male and HCB-male. Wilcoxon rank-sum test was applied.

Abrreviations: CKDB-male: urine samples provided by males with CKD; HCB-male: urine samples provided by healthy males.

**Table S3** Comparison of bacterial phylum abundance between CKDB-female and HCB-female. Wilcoxon rank-sum test was applied.

Abrreviations: CKDB-female: urine samples provided by females with CKD; HCB-female: urine samples provided by healthy females.

**Table S4** Comparison of bacterial genus abundance between CKDB-male and HCB-male. Wilcoxon rank-sum test was applied.

Abrreviations: CKDB-male: urine samples provided by males with CKD; HCB-male: urine samples provided by healthy males.

**Table S5** Comparison of bacterial genus abundance between CKDB-female and HCB-female. Wilcoxon rank-sum test was applied.

Abrreviations: CKDB-female: urine samples provided by females with CKD; HCB-female: urine samples provided by healthy females.

**Table S6** Bacteria identified using EQUC.

Abrreviations: CKDB-male: urine samples provided by males with CKD; CKDB-female: urine samples provided by females with CKD; HCB-male: urine samples provided by healthy males; HCB-female: urine samples provided by healthy females.

**Table S7** Comparisons of samples with visible cultures between HCB-male and HCB-female.Pearson Chi-square or Fisher’s exact test was used

Abrreviations: CKDB-male: urine samples provided by males with CKD; HCB-male: urine samples provided by healthy males.

**Table S8** Comparisons of samples with visible cultures between HCB-male and HCB-female.Pearson Chi-square or Fisher’s exact test was used.

Abrreviations: CKDB-female: urine samples provided by females with CKD; HCB-female: urine samples provided by healthy females.

**Table S9** Comparison of samples with visible cultures between CKDB-male and HCB-male. Pearson Chi-square or Fisher’s exact test was used.

Abrreviations: CKDB-male: bladder urine samples provided by males with CKD; HCB-male: bladder urine samples provided by healthy males.

**Table S10** Comparison of samples with visible cultures between CKDB-female and HCB-female. Pearson Chi-square or Fisher’s exact test was used.

Abrreviations: CKDB-female: bladder urine samples provided by females with CKD; HCB-female: bladder urine samples provided by healthy females.

**Figure S1.**
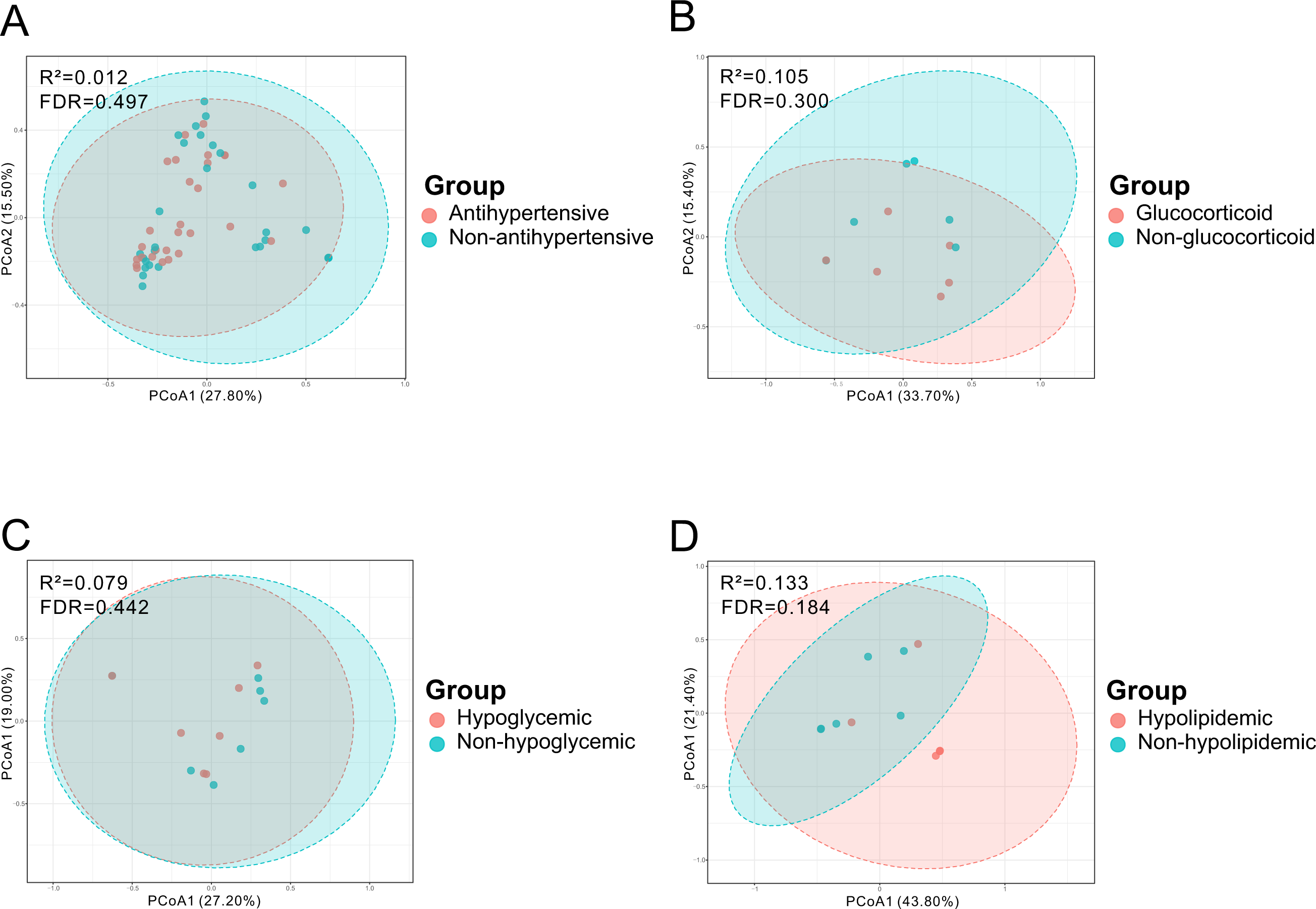
Stud design. Participants enrollment and study procedures. Abrreviations: CKDB: urine samples provided by patients with CKD; HCB: urine samples provided by healthy controls; CKDF: fecal samples provided by patients with CKD; HCF: fecal samples provided by healthy controls; CKDS: serum samples provided by CKD patient; HCS: serum samples provided by healthy controls.

**Figure S2.**
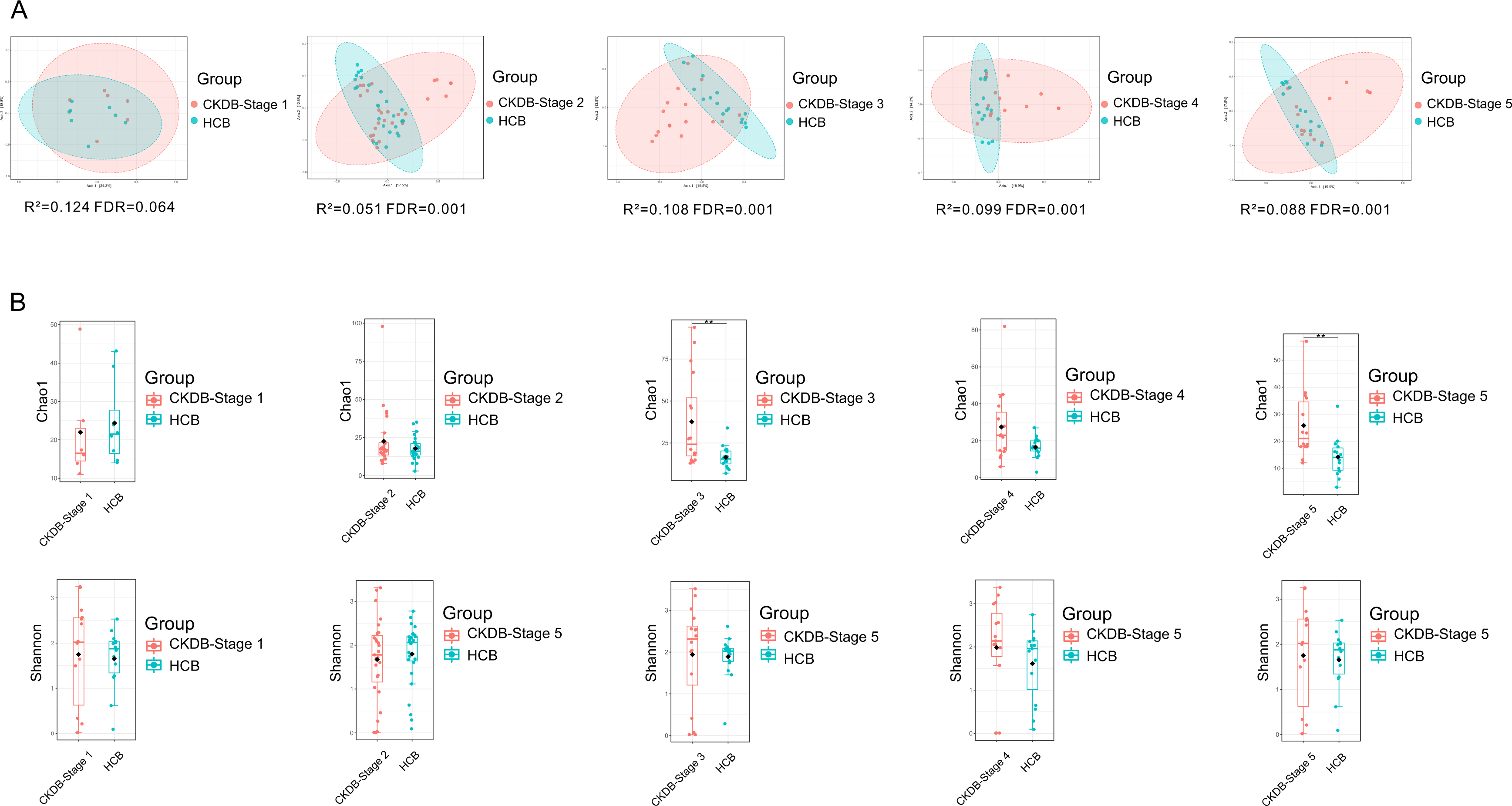
Medication usages on urinary microbiome. **(A)** Comparison of urinary microbiome between antihypertensive agent users and non-users; **(B)** Comparison of urinary microbiome between glucocorticoid agent users and non-users; **(C)** Comparison of urinary microbiome between hypoglycemic agent users and non-users; **(D)** Comparison of urinary microbiome between hypolipidemic agent users and non-users. Permutational multivariate analysis of variance (PERMANOVA) was performed for statistical comparisons of samples in the two groups. *P* value was adjusted by the Benjamini and Hochberg false discovery rate (FDR).

**Figure S3.**
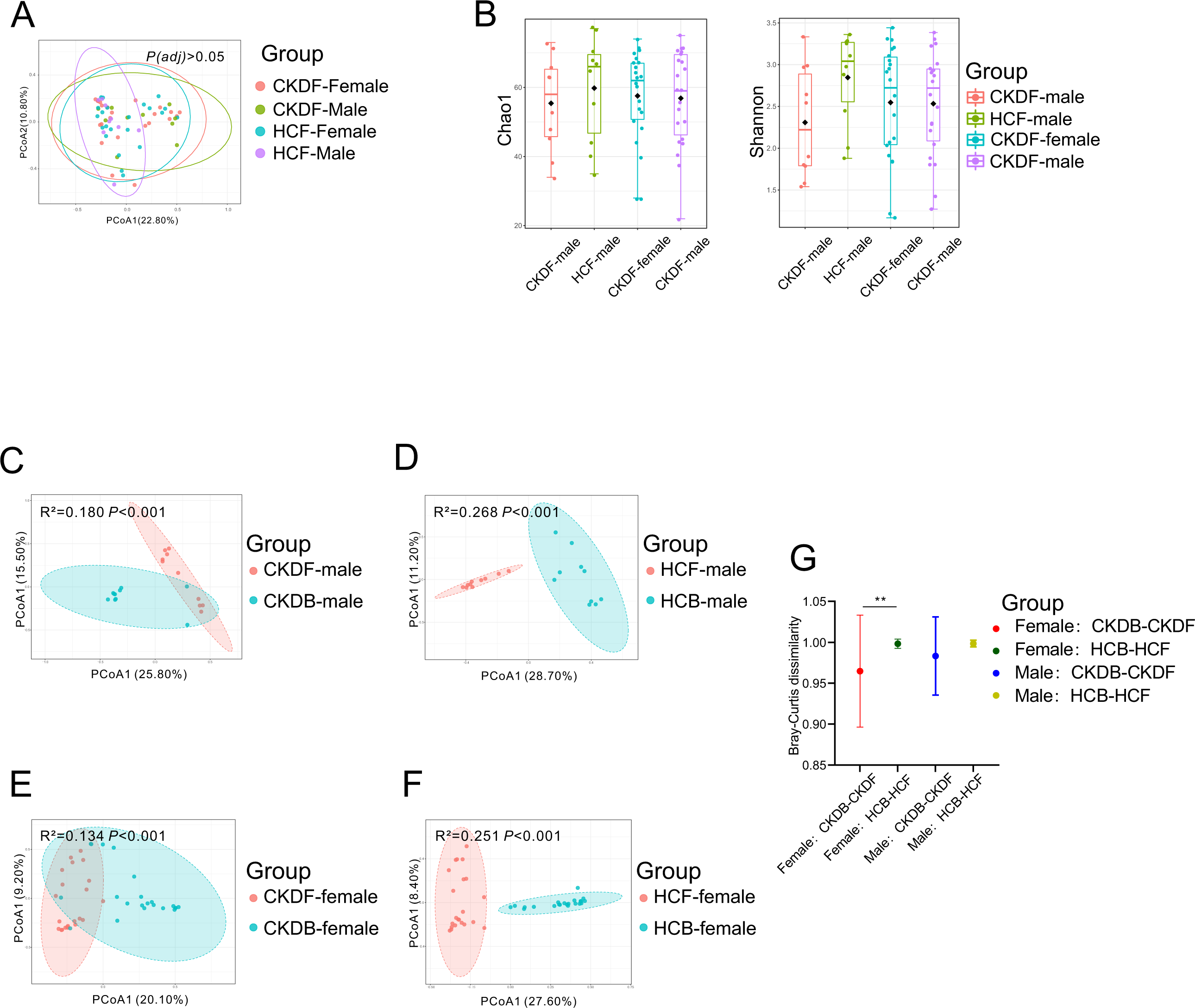
Urinary microbiome in various CKD stages. **(B)** Comparisons of microbial community between patients in CKD stage1-stage5 and controls. Permutational multivariate analysis of variance (PERMANOVA) was performed for statistical comparisons of samples in the two groups. *P* value was adjusted by the Benjamini and Hochberg false discovery rate (FDR). **(B)** Comparisons of microbial richness and diversity between patients in CKD stage1-stage5 and controls. Wilcoxon rank-sum test was performed and adjusted by Benjamini and Hochberg false discovery rate (FDR).

## Notes

### Competing Interest Statement

The authors have declared no competing interest.

### Author Declarations

The ethics committee of the Affiliated Wuxi Second Hospital of Nanjing Medical University approved the study (Ref. 2018051).

